# Resistance but not endurance training suppresses glucocorticoid-induced leucine zipper (GILZ) expression in human skeletal muscle

**DOI:** 10.1101/2024.06.14.24308924

**Authors:** Sebastian Paul, Lars Donath, Jessica Hoppstädter, Anne Hecksteden

**Affiliations:** German Sport University Cologne, Department of Training Intervention Research, 50933 Cologne, Germany; Saarland University, Department of Pharmacy, Pharmaceutical Biology, 66123 Saarbrücken, Germany; Universität of Innsbruck, Institute of Sport Science, 6020 Innsbruck, Austria; Medical University of Innsbruck, Institute of Physiology, 6020 Innsbruck, Austria

**Keywords:** GILZ, Exercise, Cardiovascular disease, Atrogenes, Gene expression

## Abstract

The glucocorticoid-induced leucine zipper (GILZ) serves as an anti-inflammatory regulator of gene expression in different tissues and is also expressed in human skeletal muscle. GILZ mediates the anti-myogenic and myotoxic side effects of statins via a shift in the Akt/FoxO signaling pathways. Recent evidence suggests that GILZ suppression is regulated by physical exercise, with external load being the decisive factor. Interestingly, statin treatment is rarely tolerated by habitually exercising individuals due to statin-associated muscle symptoms (SAMS). The opposing regulation of GILZ underpins this detrimental interaction of key measures of cardiovascular prevention. This interaction hypothetically differs between diverging exercise modalities in a mechanosensitive manner. To verify emerging evidence, we conducted a systematic search of the Gene Expression Omnibus (GEO) repository for studies reporting the acute effects of either endurance (END), conventional resistance (RT), or eccentric resistance training (ECC). 15 studies with 204 participants (22 females; 182 males, 18 to 90 years of age) were included in the analysis. Participants’ activity levels ranged from sedentary to trained. RT resulted in the highest GILZ suppression, significantly differing from the expressional change after END (-0.46 ± 1.11 vs. -0.07 ± 1.08; p = 0.03), but not from ECC (-0.46 ± 1.11 vs. -0.46 ± 0.95; p = 0.19). Furthermore, subgrouping revealed that RT-experienced participants exhibited a more pronounced GILZ suppression than their inexperienced counterparts (-0.98 ± 0.66 vs. -0.34 ± 1.16; p = 0.001). Our results strengthen the assumption that mechanical loading can be considered a key mediator of exercise-induced changes in GILZ expression.

## INTRODUCTION

The glucocorticoid-induced leucine zipper (GILZ, gene name *TSC22D3*) was first described as an immunoregulatory protein induced by dexamethasone in murine thymocytes^1^. Thereafter, GILZ expression has been reported in numerous human and murine tissues, such as the thymus, lymph nodes, bone marrow, spleen, lung, and also skeletal muscle^2–4^. Within skeletal muscle, GILZ mediates the anti-proliferative and apoptosis-inducing effects of glucocorticoids (GC)^2,4^. Similarly to glucocorticoid-mediated GILZ induction in skeletal muscle, recent evidence shows that GILZ is also elevated due to statin application^5^. Statins are a major agent for the management and treatment of hyperlipidemia and for the primary and secondary prevention of cardiovascular disease (CVD)^6–8^. Preventing cardiovascular disease continues to be of high importance as it remains the leading cause of premature mortality and rising healthcare costs, killing over four million people in Europe and over 18 million people worldwide each year^6,9^. Furthermore, the steady increase in the prevalence and mortality of cardiovascular disease between 1990 and 2019 is expected to continue, cementing the status of this problem as one of the greatest challenges in medicine^9^.

While statins effectively lower LDL-cholesterol and thus reduce the risk of CVD^6,7^, they are also associated with notable muscle-specific side effects^7^. These statin-associated muscle symptoms (SAMS) or statin myopathies range from mild myalgia^8^ to life-threatening cases of rhabdomyolysis^10^. The prevalence of SAMS is reported to vary between 5% - 29%^7,11^. Besides the acceptable benefit-to-harm ratio of statins^8^, SAMS remain the main reason for treatment discontinuation^12^. In addition, SAMS appear to be more prevalent in regularly exercising individuals and professional athletes^13,14^, which goes hand in hand with a reduced exercise-intensity tolerance^15–19^ and a reduced training adaptability in statin users^20,21^. Taken together, these findings suggest an underlying link between exercise intensity and the occurrence of SAMS. Recent research offers new insights into this presumed link, indicating that the statin-induced impairment of myogenesis is accompanied by elevated levels of GILZ expression^5^. Furthermore, the knockout of GILZ results in a resistance to statin-induced myotoxicity and to statin-induced changes in myogenin expression, an important myogenesis regulating factor (MRF). To further substantiate the involvement of GILZ in SAMS, the anti-myogenic effects of statins were mimicked by the sole overexpression of GILZ in zebrafish embryos^5^, insinuating that GILZ mediates the anti-myogenic effects of statins.

Mechanistically, SAMS are caused by a statin-induced imbalance between the protein kinase B (Akt) and the forkhead box O (FoxO) signaling pathways^5,22,23^. Akt and FoxO signaling cascades operate opposed to each other: While Akt and the mammalian target of Rapamycin (mTOR) pathway are associated with myogenesis, muscle repairment, and hypertrophy, FoxO, on the other hand, transcriptionally regulates the expression of atrophy-associated genes (atrogenes), controlling muscle protein breakdown. In the context of GILZ-mediated SAMS, three specific atrogenes are of particular interest: The muscle-specific ring finger protein-1 (MuRF-1, gene name: *TRIM63*), atrogin-1/muscle atrophy F-box (MAFbx, gene name: *FBXO32*), and Cathepsin L (CTSL, gene name: *CTSL*)^24,25^. Both MuRF-1 and MAFbx are ubiquitin E3 ligases, which are essential for protein breakdown mediated by the ubiquitin-proteasomal system (UPS)^24–27^. The UPS regulates, among others, glucocorticoid-induced muscle atrophy^27^. CTSL, on the other hand, is part of the autophagy lysosomal system (ALS)^27^ and linked to cytokine-, as well as immobilization-induced muscle atrophy^28,29^. It is noteworthy, that statins have been shown to upregulate the expression of MuRF-1, MAFbx, CTSL, and other downstream targets of FoxO^22^.

Evidence suggests that increased FoxO signaling mediates statin myopathy via an elevated expression of MuRF-1, MaFbx-1, and CTSL^22,23^. The transcriptional factor FoxO3 is particularly interesting in this context since, in addition to the expression of MuRF-1 and MaFbx-1, it also upregulates the expression of GILZ^4,5,30^.

An opposite effect to the statin-induced shift towards FoxO signaling is mediated by the peroxisome proliferator-activated receptor γ coactivator 1α (PGC1α, gene name: *PPARGC1A*), which is often described as a master regulator for endurance-associated adaptations^31,32^. PGC1α serves as a metabolic sensor of calcium signaling, which is promptly (2 h post exercise) upregulated after endurance-type exercise in rats and humans^25,31,33,34^. Sandri and coworkers observed that PGC1α transgenic mice were resistant to denervation-induced muscle atrophy and changes in muscle fiber type distribution. Moreover, the overexpression of PGC1α reduced the expression of MuRF-1, MAFbx, and CTSL by 40%^25^. These results are in line with later findings, indicating that PGC1α overexpression protects against hind limb unloading-induced muscle atrophy^32^.

Given that SAMS prevalence and habitual physical exercise appear to be linked to another^13,14^, and considering that GILZ mediates statin-induce myopathy^5^, it appears of critical importance to further investigate the relationship between physical exercise, GILZ, and SAMS on a mechanistic level. Recent evidence indicates that GILZ expression is regulated by physical exercise in a mechanosensitive manner^35^; Hecksteden et al. demonstrated that GILZ expression was acutely (30’ and 3 h post-exercise) downregulated after a single session of resistance training (RT) at an intensity of 80% 1 repetition maximum (1RM), but not after a single time-matched endurance training session (END)^35^. Thus, it is assumed that the absolute external load, as opposed to cardiovascular strain, can be regarded as the driving factor regulating exercise-induced changes in GILZ expression. Moreover, the acute downregulation of GILZ after physically demanding RT in combination with the statin-induced upregulation of GILZ may account for the harmful side effects of statins^35^. This mechanism would explain that trained individuals exercising at higher absolute loads rarely tolerate statin treatment^14^ and is further supported by the fact that physical exercise is considered a risk factor for SAMS^12,13^.

Since available exercise guidelines for CVD patients include not only moderate intense, continuous endurance exercise but also high-intensity interval training (HIIT), modified team sports, and strength training^36^, it seems reasonable to investigate the interaction between mechanical loading and GILZ expression. An improved understanding of the interaction between cardiovascular versus muscular strain and GILZ-mediated SAMS may contribute to better exercise recommendations and pharmacological strategies in CVD. Against this background, we analyzed datasets from acute exercise trials published on the Gene Expression Omnibus (GEO) repository. We hypothesized that GILZ expression is mechanosensitively affected by physical exercise.

## METHODS

### Eligibility criteria

Included data sets were required to have conducted at least one bout of either endurance (END), traditional resistance (RT), or eccentric resistance training (ECC) exercise. Studies implementing a mixture of both END and RT exercise (also called concurrent training) were excluded. Further, a sample of human skeletal muscle must have been obtained acutely (between 3-6 h) post the bout of exercise. Considered subjects were required to be described as healthy and of legal age. Studies were excluded from our analysis if the implemented exercise did not suit the requirements (i.e., concurrent exercise or insufficiently intense RT described as *rehabilitation exercise*). Subjects who were administered hormone-altering drugs (e.g., 17beta-estradiol)^37^ were excluded from the analysis. In the case that a study did conduct unilateral exertion, and did not obtain a muscle biopsy before the bout of exercise, while post-exercise, biopsies from both the trained and untrained extremity were obtained^38–40^, the data was included, and the untrained extremity was considered as pre-measurement. However, if no pre-measurement was obtained and both examined extremities did perform the same bout of exercise (i.e., but at different intensities), or one extremity did perform endurance and the other resistance type exercise, the data was excluded. Expression data were required to be obtained via either microarray or high-throughput sequencing techniques. Additionally, if the transcript of GILZ, *TSC22D3*, was not represented on the microarray or high-throughput sequencing platform, the data set was also excluded.

### Data acquisition

A systematic search of the GEO repository was conducted (Fig. 1). This search was structured according to the Preferred Reporting Items for Systematic reviews and Meta-Analyses (PRISMA) statement 2020^41^ and ended on February 29, 2024. The search strings used are documented in the supplementary materials (supp. 2.1.1). Screening was conducted manually and structured according to the aforementioned inclusion criteria. In the rare case where multiple microarray probes were assigned to only one RefSeq ID of interest (a total of five occurrences), all corresponding probe signals were averaged. If one RefSeq ID was represented by multiple probes, and the probes were denoted with versions, the signal of the latest probe version was considered (one occurrence). If included studies obtained multiple muscle biopsies within our determined time frame, the biopsy closest to 6 h post-exercise was chosen. This decision was based on recent evidence demonstrating that translation- and transcription-initiation factor expression, as well as global gene expression, can be expected to peak within the latter phase of the selected time span^42–44^.

**Figure 1.**
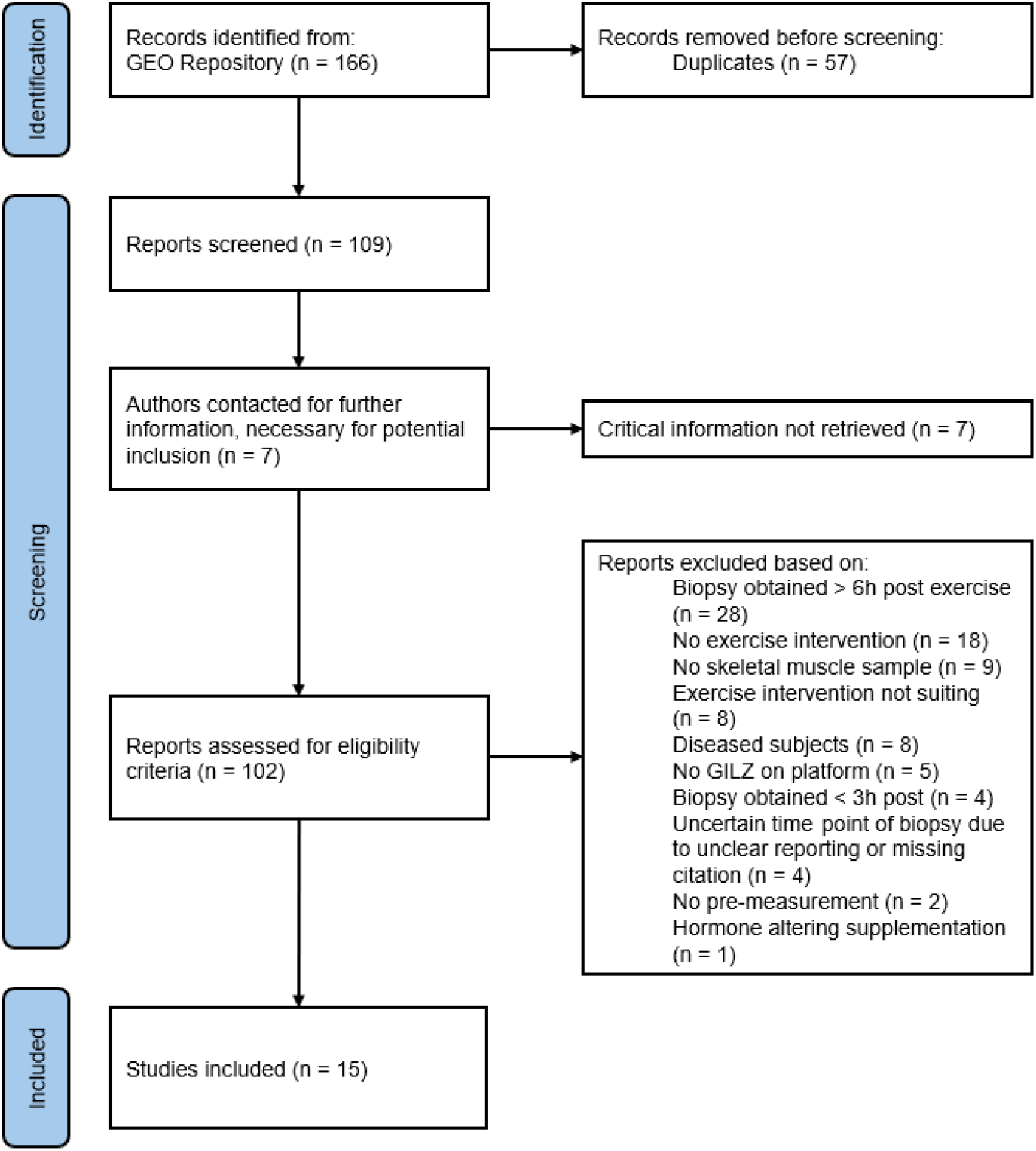
Flow chart of the systematic search process in accordance with the Preferred Reporting Items for Systematic Reviews and Meta-Analyses (PRISMA) 2020 statement^41^.

If subjects were tested multiple times, for example, before and after an exercise intervention lasting several weeks^40,45^, or after a high-load and a low-load exercise bout of RT^46^, measurements of only one of the occasions were analyzed. Thereby we intended to avoid multiple inclusions of identical subjects. In these cases, the measurement after the exercise intervention and acutely after the high-load RT session were included in the analysis. This decision was based on the assumption that higher external loads are apparent post higher relative exercise intensities and post familiarization, ultimately resulting in a more pronounced suppression of GILZ expression. Additionally, in one particular placebo-controlled study, half of the subjects were assigned to a 17beta-estradiol treatment, lasting for eight days^37^. In this case, only the placebo group was included in our analysis.

### Data processing

Data analysis and visualization were performed using R Statistical Software^47^ in the R Studio IDE^48^. To account for variation between trials, within-trial robust scaling of the raw data was performed. Robust scaling is considered a non-parametric alternative to the z-score and is calculated by dividing the difference between the respective value and the median by the interquartile range. Consequently, the scaled values represent the difference between the current value and the individual ‘baseline’ in units of interquartile ranges. Given that gene expressional data generally cannot be assumed to be distributed normally^49^, and varies substantially on inter- and intraindividual level^50,51^, this scaling method seemed appropriate. Subsequently, changes in gene expression from pre-to post-exercise were calculated (Δ). Datapoints deviating more than three times the interquartile range from the median were considered outliers and excluded from the analysis.

### Statistical analysis

Gene expression data is robust scaled and is presented in arbitrary units (AU) as mean ± standard deviation. Normal distribution of the robustly scaled values was assessed visually with the help of quantile-quantile plots. ANOVA was computed to compare changes in Δ gene expression between groups. Independent Welch’s t-test was used to compare ΔGILZ expression between the subset of trained and untrained individuals within the RT group. Critical threshold of significance was set at p < 0.05 and is denoted with asterisks (p < 0.05: *; p < 0.01: **; p < 0.001: ***).

## RESULTS

A total of 15 studies and 204 participants (22 female; 182 male; see Tab. 1) were included in the analysis. Six studies conducted END, five RT, two ECC, and two split their subjects into either END or RT. Three of the seven RT protocols investigated RT-trained or familiarized participants^46,52,53^ (Tab. 1). Participant’s habitual activity level ranged from sedentary to endurance and strength trained (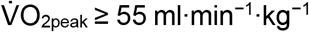, and barbell back squat ≥ 1.5 times bodyweight), and the age of examined individuals ranged from 18 to 90 years (Tab. 1). A summary of included studies, implemented training regimes, and participants’ characteristics is provided in table 1 (Tab.1).

**Table 1:** Summary of participants’ characteristics and implemented exercise regimes of included trials. Age and BMI are presented either in mean ± SD, median and IQR, or as a range.

| Author, Publication date | GEO Accession | n | Age [years] | BMI [kg/m<sup>2</sup>] | Exercise mode | Exercise selection | Intensity | Volume | time point biopsy | Training status |
| --- | --- | --- | --- | --- | --- | --- | --- | --- | --- | --- |
| **Endurance trials** |  |  |  |  |  |  |  |  |  |  |
| Dickinson et al., 2018 | GSE107934 | 6 (m) | 27 ± 3 | NA | END | Cycle ergometer | 70% max HR | 40 min | 4 h post | Recreationally active |
| Makhnovskii et al., 2022 | GSE164081 | 10 (m) | 32, (IQR: 30-36) | NA | END | Cycle ergometer | 50&100% Power @ La 4mmol/L | 60 min | 6 h post | Endurance trained ($\dot{V}O_{2max}$: 54-60 ml·min<sup>-1</sup>·kg<sup>-1</sup>) |
| Neubauer et al., 2013 | GSE43856 | 8 (m) | 25 ± 4.1 | NA | END | Cycle ergometer & Treadmill running | 105% PPO of gas exchange threshold & self-paced 10km trial speed | 120 min | 4 h post | Endurance trained ($\dot{V}O_{2max}$: 56.3 ± 6.7 ml·min<sup>-1</sup>·kg<sup>-1</sup>) |
| Popov et al., 2018 | GSE86931 | 2 (m) | 23 (IQR: 20-27) | NA | END | Cycle ergometer | 70% $\dot{V}O_{2max}$ | 60 min | 4 h post | Endurance trained ($\dot{V}O_{2max}$: 61 ml·min<sup>-1</sup>·kg<sup>-1</sup>) |
| Popov et al., 2019<sup>†</sup> | GSE120862 | 7 (m) | 21 (IQR: 21-24) | 23 (IQR: 23-24) | END | Modified cycle ergometer | 65% PPO ramp test | 60 min | 4 h post | Sedentary |
| Rowlands et al., 2011 | GSE27285 | 8 (m) | 32.8 ± 6.4 | NA | END | Cycle ergometer | 50-90% PPO | 105 min | 3 h post | Endurance trained subjects ($\dot{V}O_{2max}$: 59.97 ml·min<sup>-1</sup>·kg<sup>-1</sup>) |
| Rubenstein et al., 2022 | GSE151066 | 19 (4f \| 15m) | 69.5 ± 3.8 | 26.5 ± 3.8 | END | Cycle ergometer | 70% HRR | 40 min | 3 h post | Active vs. Inactive |
| Vissing et al., 2018 | GSE59088 | 7 (m) | 23.4 ± 0.8 | NA | END | Cycle ergometer | 60% $\dot{V}O_{2peak}$ | 120 min | 5 h post | 10 weeks of familiarization |
| **Resistance training trials** |  |  |  |  |  |  |  |  |  |  |
| Centner et al., 2022 | GSE195585 | 30 (m) | 24.7 ± 2.5 | 22.8 ± 2.4 | RT | Knee extension machine | 80% 1RM | 40 reps | 4 h post | Recreationally active |
| Dickinson et al., 2018 | GSE107934 | 6 (m) | 27 ± 3 | NA | RT | Isotonic unilateral leg extension | 65% 1RM | 80 reps | 4 h post | Recreationally active |
| Liu et al., 2010 | GSE24235 | 7 (4f \| 3m) | 23.9 ± 2.12 | 23.9 ± 4.5 | RT | Variation of elbow flexing and extending exercises | 6RM | 90 reps | 4 h post | 12 weeks of familiarization |
| McIntosh et al., 2023 | GSE220899 | 11 (m) | 23 ± 4 | 27 ± 3 | RT | Squats & knee extension machine | 80% 1RM to failure | to failure | 6 h post | Trained (BB squat ≥ 1.5 times BW) |
| Raue et al., 2012 | GSE28422 | 12 (6f \| 6m) | 84 ± 3 | 26 ± 3 | RT | Knee extension machine | 70-75% 1RM | 30 reps | 4 h post | 12 weeks of familiarization |
| Raue et al., 2012 | GSE28422 | 16 (8f \| 8m) | 24 ± 4 | 25 ± 5 | RT | Knee extension machine | 70-75% 1RM | 30 reps | 4 h post | 12 weeks of familiarization |
| Vissing et al., 2018 | GSE59088 | 7 (m) | 23.4 ± 0.8 | NA | RT | Leg press, knee extension, and hamstring curl | 12RM | 144 reps | 5 h post | 10 weeks of familiarization |
| Zambon et al., 2003<sup>†</sup> | GSE1832 | 4 (m) | btw. 31-51 | NA | RT | Unilateral isotonic knee extension | 80% 1RM | 80 reps | 6 h post | Sedentary |

| Eccentric resistance trials |  |  |  |  |  |  |  |  |  |  |
| --- | --- | --- | --- | --- | --- | --- | --- | --- | --- | --- |
| Hyldahl et al., 2011<sup>†</sup> | GSE23697 | 35 (m) | 20.9 ± 0.5 | NA | ECC | Biodex isokinetic dynamometer | Max. effort | 100 reps | 3 h post | Sedentary |
| MacNeil et al., 2010<sup>‡</sup> | GSE19062 | 9 (m) | 21 ± 2 | NA | ECC | Biodex isokinetic dynamometer | Max. effort | 150 reps | 3 h post | Sedentary |

<sup>†</sup> No pre-measurement, unilateral exercise; unexercised leg was considered as pre. <sup>‡</sup> 17beta-estradiol treatment, only control subjects were included. BMI: Body-mass-index. min: minutes. reps: repetitions. btw.: between. HRR: Heart rate reserve. PPO: Peak power output. BB squat: Barbell back squat. BW: bodyweight.

### GILZ

All three exercise regimes did result in a negative expressional change of GILZ: While END reduced GILZ expression only marginally (Δ = -0.07 ± 1.08), the suppression of GILZ following RT and ECC was more pronounced (Δ = -0.46 ± 1.11 and -0.46 ± 0.95). The magnitude of Δ GILZ expression differed significantly between END and RT (p = 0.03) but not between RT and ECC (p = 0.19), nor between END and ECC (p = 0.07) (Fig. 2). When subgrouping RT into trained participants versus novices, trained individuals experienced significantly greater reductions in GILZ expression compared to the novices (p = 0001; Δ = -0.98 ± 0.66 vs. -0.34 ± 1.16) (Fig. 3).

**Figure 2.**
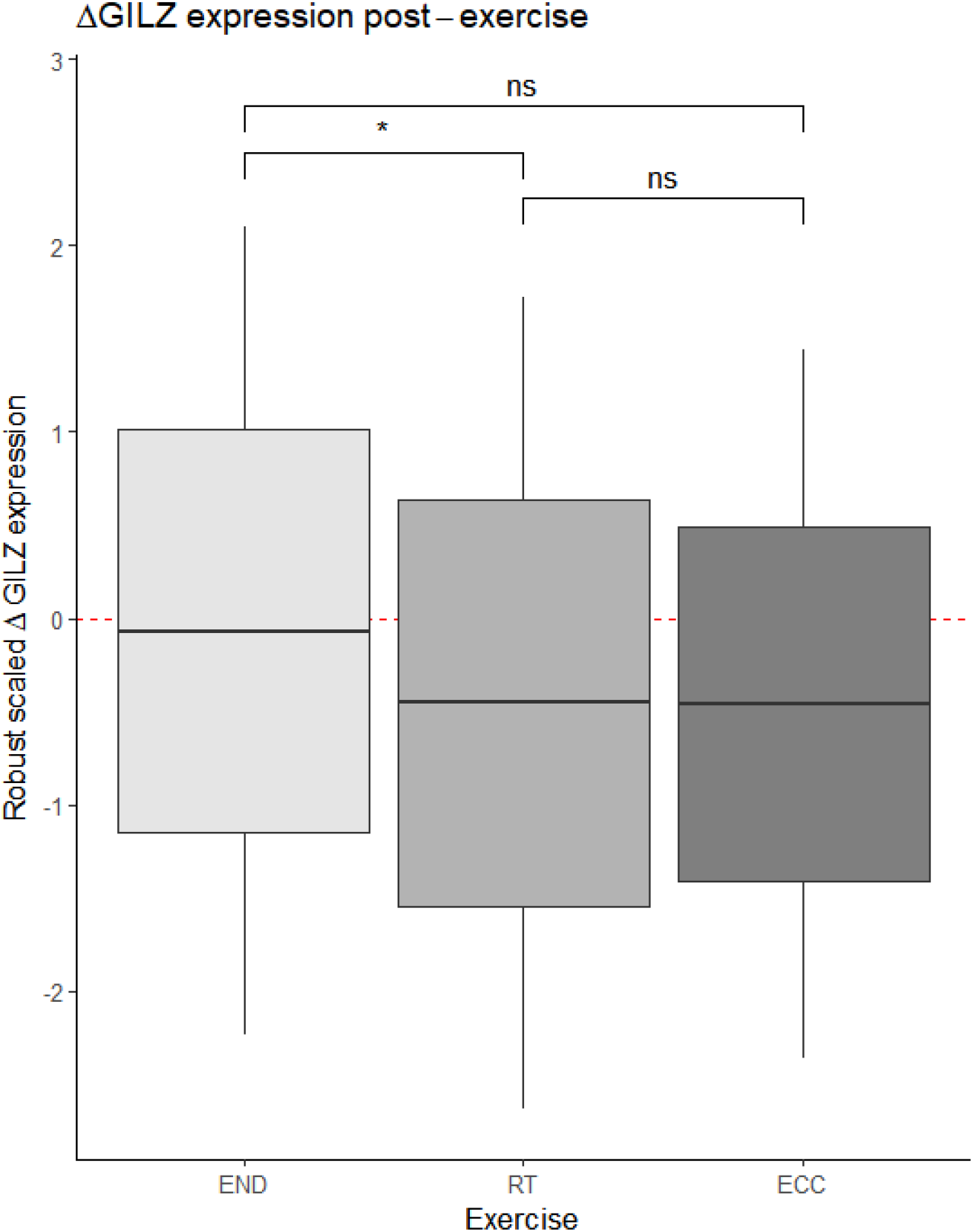
Group comparison of ΔGILZ expression post different exercise modalities. Black line depicts the mean, outer lines: SD, and whiskers: twofold SD. Red dashed zero-line indicates the reference for no expression change from pre- to post-exercise. *p < .05, **p < .01, ***p < 0.001, ns: non-significant.

**Figure 3.**
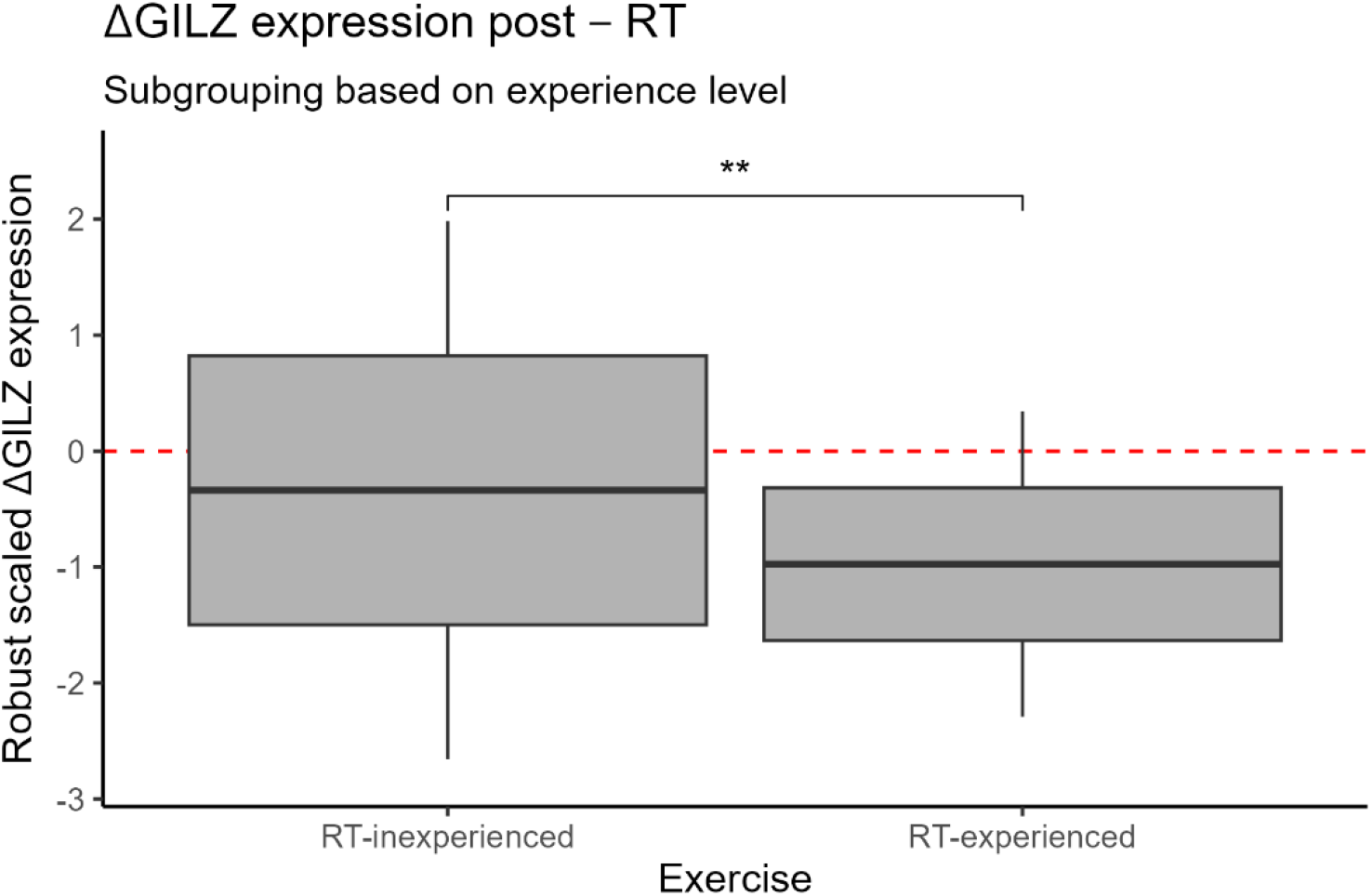
ΔGILZ expression post-RT, sample size subdivided into RT-experienced (n = 53) and - inexperienced subjects (n = 40). The black line depicts the mean, outer lines: SD, and whiskers: twofold SD. Red dashed zero- line indicates the reference for no expressional change from pre to post exercise. *p < .05, **p < .01, ***p < 0.001, ns: non-significant.

### Atrogenes

Only ECC training resulted in a negative ΔMuRF1 (Δ = -0.4 ± 1.04) expression. RT and END resulted in a positive ΔMuRF1 expression (Δ = 0.46 ± 1.08; 0.59 ± 1.08). The ΔMuRF1 expression after ECC differed significantly from both RT (p < 0.0001; Δ = -0.4 ± 1.04 vs. 0.46 ± 1.08) and END (p < 0.0001; Δ = -0.4 ± 1.04 vs. 0.59 ± 1.08) (Fig. 4A). Concerning ΔMAFbx expression, END resulted in a positive expression change (Δ = 0.47 ± 1.07), while both RT and ECC led to a notable suppression of MAFbx expression (Δ = -0.54 ± 0.81 and -0.65 ± 0.74). ΔMAFbx expression of RT and ECC differ significantly from END (p < 0.0001; p < 0.0001; Δ = 0.47 ± 1.07 vs. -0.54 ± 0.81, and 0.47 ± 1.07 vs. -0.65 ± 0.74). No significant difference between RT and ECC groups was found (p = 0.24; Δ = -0.54 ± 0.81 vs. -0.65 ± 0.74) (see Fig. 4B). Furthermore, both END and ECC resulted in a CTSL upregulation (Δ = 0.64 ± 0.87; 0.24 ± 1.09), while RT caused CTSL downregulation (Δ = -0.03 ± 1.1). Significant group differences in ΔCTSL expression were found for END versus RT (p < 0.0001; Δ = 0.64 ± 0.87 vs. -0.03 ± 1.1) and END versus ECC (p = 0.041; Δ = 0.64 ± 0.87 vs. 0.24 ± 1.09), but not when comparing RT to ECC (p = 0.18; Δ = -0.03 ± 1.1 vs. 0.24 ± 1.09) (Fig. 4C).

**Figure 4.**
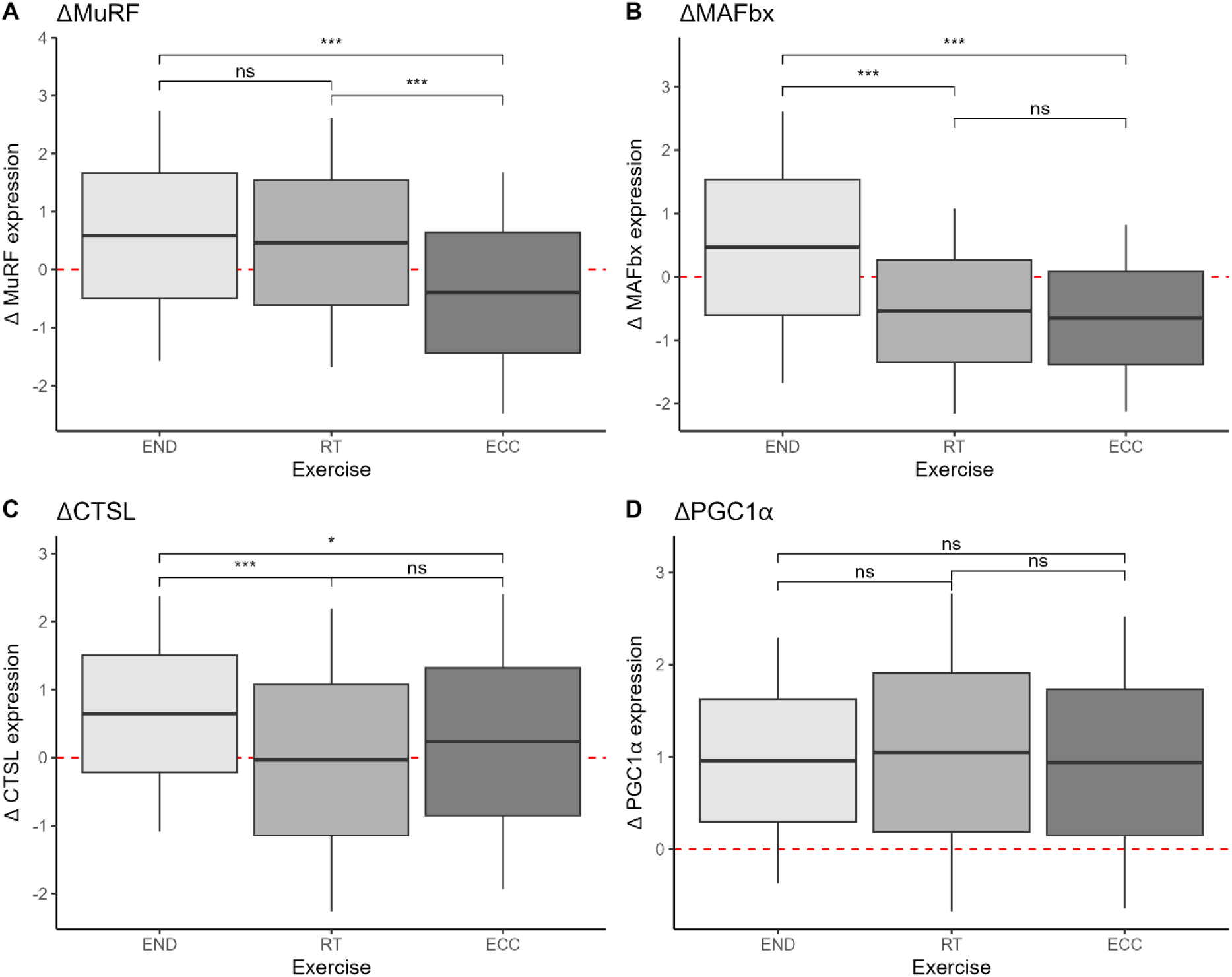
**A-D** Group comparison of ΔAtrogenes and ΔPGC1α expression post different exercise modalities. Black line depicts the mean, outer lines: SD, and whiskers: twofold SD. Red dashed zero-line indicates the reference for no expressional change from pre to post exercise. *p < .05, **p < .01, ***p < 0.001, ns: non-significant.

### PGC1α

All exercise modalities resulted in an upregulation of PGC1α expression (END: Δ = +0.61 ± 0.665; RT: Δ = +1.01 ± 0.713; ECC: Δ = +0.94 ± 0.79). No significant between-group differences for PGC1α expression were observed (p > 0.05) (see Fig. 4D).

## DISCUSSION

This is the first study that systematically screened data from the GEO repository to investigate the effect of different training modalities on GILZ expression. We assumed that physical exercise affects GILZ expression and hypothesized that concentric and eccentric strength training would lead to a more pronounced GILZ suppression than endurance training. Our results indicate notably suppressed GILZ expression after traditional and eccentric RT. Furthermore, the GILZ expression post RT differed significantly from the GILZ espression following endurance training (Fig. 2). This finding supports our hypothesis that GILZ expression is influenced by physical exercise, particularly mechanical loading. Based on a comparatively small sample (Tab. 1) and, consequently, diminished study power, changes in GILZ expression after eccentric training did not significantly differ from endurance training. Although insignificant, eccentric training resulted in the downregulation of GILZ, which was more than six-fold greater than post-endurance training.

Interestingly, the participants’ training status seems to play a critical role in the magnitude of GILZ suppression post-exercise. We found that RT-trained participants experienced a more significant GILZ suppression compared to their untrained counterparts (Fig. 3). This finding further strengthens our hypothesis that absolute external load is the main determinant influencing GILZ expression post-physical exercise since resistance-trained participants performed the same relative intensity in an RT session with absolutely higher external loads, as compared to strength training inexperienced individuals^54^. This assumption is in line with the findings from Hecksteden and colleagues, demonstrating a significant downregulation of GILZ expression acutely after resistance training, only in RT-trained, but not in strength training inexperienced participants^35^. It thus seems that absolute mechanical external load is the main driver of exercise-induced GILZ expression changes. This interaction may explain why exercising individuals rarely tolerate statin therapy and further suggests that regular strength-related exercise may increase the risk of statin myopathy^12–14^.

When examining post-exercise atrogene expression, it appears that mechanically demanding exercise regimes generally tend to suppress MuRF1, MAFbx, and CTSL expression, which again, stands in contrast to the statin-induced elevation of FoxO downstream targets ^22,23^ and the heightened expression levels post-END (Fig. 4 A-C). More specifically, our results show that a single bout of endurance exercise results in a positive change of MuRF1, MAFbx, and CTSL expression (ΔMuRF1 = 0.59 ± 1.08; ΔMAFbx = 0.47 ± 1.07; ΔCTSL = 0.64 ± 0.87). This observation is in line with findings from Louis and Coffey et al., demonstrating that both running (30 minutes at 75%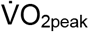), and cycling (60 min at 70%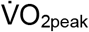) induced an acute (1 to 4 h post-exercise) upregulation of MuRF1 and MAFbx^43,55^. Similarly, Schwalm and colleagues found an immediate (1 h post) upregulation of CTSL after 2 h of cycling at an intensity of 70%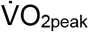. Interestingly, 2 h of cycling at lower intensities (55%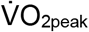) did not significantly change CTSL expression^56^. Thus, an intensity-dependent CTSL expression seems likely.

Our observations expand on these findings, showing that only mechanically demanding eccentric resistance exercise led to a meaningful MuRF1 suppression, which significantly differed from the positive ΔMuRF1 expression after END and traditional RT (Fig. 4A). These findings are at least partly in line with available evidence, suggesting that RT, like END, acutely upregulates MuRF1 expression. It appears that moderately intense (65-70% of 1RM) RT, consisting of only 30 repetitions of knee extensions, is a sufficient stimulus to acutely (1 to 4 h post-exercise) upregulate MuRF1^43,57^. Further, Nedegaard et al. reported an upregulation of MuRF1 3 h post 300 concentric contractions (i.e., unilateral box step-ups), while the MuRF1 expression in the contralateral leg, performing the eccentric contractions (i.e., stepping down from the box), remained unchanged^58^. Compelling evidence on MuRF1 expression post-ECC is still lacking as gene expression data post eccentric resistance exercise are scarce. Considering the available evidence, it might be reasonable to assume that traditional RT acutely upregulates MuRF1 expression, while exercise implementing higher external loads (potentially mediated via eccentric contractions) leads to no acute change in MuRF1 expression or even to a slight suppression.

Our results show that both RT and ECC induce an acute downregulation of MAFbx, which differs significantly from the upregulation of MAFbx expression following END (Fig. 4B). The majority of published articles did not observe changes in MAFbx expression acutely (1 to 4 h) after RT^43,44,55,57^. In contrast to these findings, when expanding the time frame after exercise cessation (5-12h post), Louis et al. did find a significant suppression in MAFbx expression after moderately intense (70% of 1RM, 30 reps) resistance exercise^43^. In this regard, Stefanetti and colleagues also observed a decreased MAFbx expression 2.5 h post resistance exercise, which continued to decrease to ultimately reach its minimum at 5 h post-RT^59^. Consequently, MAFbx expression seems acutely (1-4 h post) unchanged after RT, and then, over the time course of 5-12 h post-exercise, decreases to levels below the baseline values. Our results strengthen the assumption that changes in MAFbx expression are heavily time-dependent (Tab.1 & Fig. 4B). Taking existing evidence and our results together, it appears that RT-induced downregulation of MAFbx can be expected to occur within a time frame of 4-12 h after resistance exercise. Thus, it might be assumed that not only the opposing regulation of GILZ, but also the opposing regulation of MuRF1, MAFbx, and CTSL, post-mechanically demanding exercise and statin application, might lead to elevated risk of SAMS occurrence in habitually exercising individuals.

All considered exercise modalities (i.e., END, RT, and ECC) led to a positive change in PGC1α expression of a similar magnitude, with no significant between-group differences. Since PGC1α is considered an important sensor for motor neuron-induced calcium signaling^25^, it seems reasonable that all exercises containing sufficient motor neuron-induced muscle contractions would lead to a PGC1α upregulation. This assumption is supported by the fact that metabolic perturbations, such as intracellular Ca^++^ oscillation, acutely upregulate PGC1α expression and activity^60^, in both animal and human models^33,61^. Moreover, the total number of consecutive muscle contractions may be essential in PGC1α upregulation and, subsequently, the PGC1α-mediated suppression of FoxO signaling^62^. Although PGC1α expression is known to be upregulated by a low-amplitude and low-frequency Ca^++^ oscillation, typically induced by high-volume and low-intensity endurance exercise^60^, recent evidence shows that PGC1α expression is also upregulated post resistance type exercises^63,64^. Specifically, resistance exercise seems to upregulate the expression of a particular PGC1α splice variant called PGC1α4^64^. This splice variant ultimately promotes muscle fiber hypertrophy and enhances anaerobic glycolysis^63,64^.

Our retrospective analysis of GEO datasets comprises limitations that need to be addressed: A substantial amount of between-trial heterogeneity regarding participants’ characteristics and performed exercise regimes needs to be stated. This applies to the training status of the examined individuals, which affects absolute external loads during RT sessions and the resulting implemented absolute exercise intensities. Further, only four^45,46,52,53^ out of seven RT trials^39,65,66^ (∼57.1%) examined trained or familiarized subjects. This discrepancy in familiarization level may affect the magnitude of RT-induced changes in GILZ expression when RT trials are examined collectively. The same limitation holds true for the included END trials, where five^53,67–70^ out of eight^40,66,71^ (62.5%) studies examined endurance-trained or familiarized subjects. Furthermore, the intensity of the exercises implemented in the examined trials varied substantially. For example, two END trials investigated gene expression changes in endurance-trained males post high-intensity interval training (HIIT)^67,69^. In both of these studies, subjects performed exercise bouts of maximal (100%) or close to maximal (90%) intensities (peak power output; PPO) interspersed by active recovery segments at lower intensities (50% PPO). Our analysis revealed that these particular exercise trials resulted in a downregulation of GILZ that was exceptionally high compared to the other END trials, which involved less intense and continuous endurance exercise (results not shown). Nonetheless, our findings clearly demonstrate a significant downregulation of GILZ expression post-RT, specifically when subjects are familiarized and thus perform the same relative intensity at absolute higher external loads. These findings have been obtained despite the aforementioned heterogeneity between implemented exercise regimes and the training status of included studies.

## CONCLUSION

Our results strongly support the hypothesis that particularly mechanical loading serves as a key mediator of training-induced suppression of GILZ acutely post exercise. Notably, this result has been obtained despite the substantial differences in age, level of habitual exercise, and exercise dose across studies. If confirmed, these findings may contribute to a more harmonized treatment of CVD, consisting of both statin medication and specifically adapted exercise training. To further clarify the underlying relationship between exercise, GILZ expression and SAMS, further research is needed that considers the use of statins and different exercise modalities.

## Supporting information

Supplemental Data Acquisition

## Data availability statement

The data were obtained from the publicly available GEO repository [https://www.ncbi.nlm.nih.gov/gds], included trials are referenced with their respective GEO accession number in Table 1.

## Authors’ contributions

A. Hecksteden, L. Donath, and J. Hoppstädter conceptualized the study.

S. Paul performed the search for trials, screening, data extraction, analysis, and visualization of the data.

J. Hoppstädter assisted in the data extraction.

All authors interpreted the results.

S. Paul wrote the manuscript with assistance from L. Donath and A. Hecksteden.

All authors provided feedback and final approval of the manuscript.

## REFERENCES

1. D’Adamio F, Zollo O, Moraca R, et al. A New Dexamethasone-Induced Gene of the Leucine Zipper Family Protects T Lymphocytes from TCR/CD3-Activated Cell Death. Immunity. 1997;7(6):803–812. doi:10.1016/S1074-7613(00)80398-2

2. Bruscoli S, Donato V, Velardi E, et al. Glucocorticoid-induced Leucine Zipper (GILZ) and Long GILZ Inhibit Myogenic Differentiation and Mediate Anti-myogenic Effects of Glucocorticoids*. Journal of Biological Chemistry. 2010;285(14):10385–10396. doi:10.1074/jbc.M109.070136

3. Cannarile L, Zollo O, D’Adamio F, et al. Cloning, chromosomal assignment and tissue distribution of human GILZ, a glucocorticoid hormone-induced gene. Cell Death Differ. 2001;8(2):201–203. doi:10.1038/sj.cdd.4400798

4. Bruscoli S, Riccardi C, Ronchetti S. GILZ as a Regulator of Cell Fate and Inflammation. Cells. 2021;11(1):122. doi:10.3390/cells11010122

5. Hoppstädter J, Valbuena Perez JV, Linnenberger R, et al. The glucocorticoid-induced leucine zipper mediates statin-induced muscle damage. The FASEB Journal. 2020;34(3):4684–4701. doi:10.1096/fj.201902557RRR

6. Catapano AL, Graham I, De Backer G, et al. 2016 ESC/EAS Guidelines for the Management of Dyslipidaemias. Eur Heart J. 2016;37(39):2999–3058. doi:10.1093/eurheartj/ehw272

7. Grundy SM, Stone NJ, Bailey AL, et al. 2018 AHA/ACC/AACVPR/AAPA/ABC/ACPM/ADA/AGS/APhA/ASPC/NLA/PCNA Guideline on the Management of Blood Cholesterol: Executive Summary: A Report of the American College of Cardiology/American Heart Association Task Force on Clinical Practice Guidelines. J Am Coll Cardiol. 2019;73(24):3168–3209. doi:10.1016/j.jacc.2018.11.002

8. Cai T, Abel L, Langford O, et al. Associations between statins and adverse events in primary prevention of cardiovascular disease: systematic review with pairwise, network, and dose-response meta-analyses. BMJ. 2021;374:n1537. doi:10.1136/bmj.n1537

9. Roth GA, Mensah GA, Johnson CO, et al. Global Burden of Cardiovascular Diseases and Risk Factors, 1990–2019. J Am Coll Cardiol. 2020;76(25):2982–3021. doi:10.1016/j.jacc.2020.11.010

10. Moßhammer D, Schaeffeler E, Schwab M, Mörike K. Mechanisms and assessment of statin-related muscular adverse effects. Br J Clin Pharmacol. 2014;78(3):454–466. doi:10.1111/bcp.12360

11. Stroes ES, Thompson PD, Corsini A, et al. Statin-associated muscle symptoms: impact on statin therapy-European Atherosclerosis Society Consensus Panel Statement on Assessment, Aetiology and Management. Eur Heart J. 2015;36(17):1012–1022. doi:10.1093/eurheartj/ehv043

12. Laufs U, Filipiak KJ, Gouni-Berthold I, Catapano AL, SAMS expert working group. Practical aspects in the management of statin-associated muscle symptoms (SAMS). Atheroscler Suppl. 2017;26:45–55. doi:10.1016/S1567-5688(17)30024-7

13. Dirks AJ, Jones KM. Statin-induced apoptosis and skeletal myopathy. Am J Physiol Cell Physiol. 2006;291(6):C1208–1212. doi:10.1152/ajpcell.00226.2006

14. Sinzinger H, O’Grady J. Professional athletes suffering from familial hypercholesterolaemia rarely tolerate statin treatment because of muscular problems. Br J Clin Pharmacol. 2004;57(4):525–528. doi:10.1111/j.1365-2125.2003.02044.x

15. Kearns AK, Bilbie CL, Clarkson PM, et al. The creatine kinase response to eccentric exercise with atorvastatin 10 mg or 80 mg. Atherosclerosis. 2008;200(1):121–125. doi:10.1016/j.atherosclerosis.2007.12.029

16. Parker BA, Augeri AL, Capizzi JA, et al. Effect of statins on creatine kinase levels before and after a marathon run. Am J Cardiol. 2012;109(2):282–287. doi:10.1016/j.amjcard.2011.08.045

17. Thompson PD, Nugent AM, Herbert PN. Increases in creatine kinase after exercise in patients treated with HMG Co-A reductase inhibitors. JAMA. 1990;264(23):2992.

18. Thompson PD, Gadaleta PA, Yurgalevitch S, Cullinane E, Herbert PN. Effects of exercise and lovastatin on serum creatine kinase activity. Metabolism. 1991;40(12):1333–1336. doi:10.1016/0026-0495(91)90039-y

19. Thompson PD, Zmuda JM, Domalik LJ, Zimet RJ, Staggers J, Guyton JR. Lovastatin increases exercise-induced skeletal muscle injury. Metabolism. 1997;46(10):1206–1210. doi:10.1016/s0026-0495(97)90218-3

20. Mikus CR, Boyle LJ, Borengasser SJ, et al. Simvastatin impairs exercise training adaptations. J Am Coll Cardiol. 2013;62(8):709–714. doi:10.1016/j.jacc.2013.02.074

21. Morales-Palomo F, Ramirez-Jimenez M, Ortega JF, Moreno-Cabañas A, Mora-Rodriguez R. Exercise Training Adaptations in Metabolic Syndrome Individuals on Chronic Statin Treatment. J Clin Endocrinol Metab. 2020;105(4):dgz304. doi:10.1210/clinem/dgz304

22. Mallinson JE, Constantin-Teodosiu D, Sidaway J, Westwood FR, Greenhaff PL. Blunted Akt/FOXO signalling and activation of genes controlling atrophy and fuel use in statin myopathy. J Physiol. 2009;587(Pt 1):219–230. doi:10.1113/jphysiol.2008.164699

23. Hanai J ichi, Cao P, Tanksale P, et al. The muscle-specific ubiquitin ligase atrogin-1/MAFbx mediates statin-induced muscle toxicity. J Clin Invest. 2007;117(12):3940–3951. doi:10.1172/JCI32741

24. Sandri M, Sandri C, Gilbert A, et al. Foxo transcription factors induce the atrophy-related ubiquitin ligase atrogin-1 and cause skeletal muscle atrophy. Cell. 2004;117(3):399–412. doi:10.1016/s0092-8674(04)00400-3

25. Sandri M, Lin J, Handschin C, et al. PGC-1α protects skeletal muscle from atrophy by suppressing FoxO3 action and atrophy-specific gene transcription. Proc Natl Acad Sci USA. 2006;103(44):16260–16265. doi:10.1073/pnas.0607795103

26. Stitt TN, Drujan D, Clarke BA, et al. The IGF-1/PI3K/Akt pathway prevents expression of muscle atrophy-induced ubiquitin ligases by inhibiting FOXO transcription factors. Mol Cell. 2004;14(3):395–403. doi:10.1016/s1097-2765(04)00211-4

27. Wilburn D, Ismaeel A, Machek S, Fletcher E, Koutakis P. Shared and distinct mechanisms of skeletal muscle atrophy: A narrative review. Ageing Research Reviews. 2021;71:101463. doi:10.1016/j.arr.2021.101463

28. Ebisui C, Tsujinaka T, Morimoto T, et al. Interleukin-6 Induces Proteolysis by Activating Intracellular Proteases (Cathepsins B and L, Proteasome) in C2C12 Myotubes. Clinical Science. 1995;89(4):431–439. doi:10.1042/cs0890431

29. Bialek P, Morris C, Parkington J, et al. Distinct protein degradation profiles are induced by different disuse models of skeletal muscle atrophy. Physiol Genomics. 2011;43(19):1075–1086. doi:10.1152/physiolgenomics.00247.2010

30. Asselin-Labat ML, David M, Biola-Vidamment A, et al. GILZ, a new target for the transcription factor FoxO3, protects T lymphocytes from interleukin-2 withdrawal–induced apoptosis. Blood. 2004;104(1):215–223. doi:10.1182/blood-2003-12-4295

31. Lundby C, Jacobs RA. Adaptations of skeletal muscle mitochondria to exercise training. Experimental Physiology. 2016;101(1):17–22. doi:10.1113/EP085319

32. Cannavino J, Brocca L, Sandri M, Bottinelli R, Pellegrino MA. PGC1-α over-expression prevents metabolic alterations and soleus muscle atrophy in hindlimb unloaded mice. J Physiol. 2014;592(20):4575–4589. doi:10.1113/jphysiol.2014.275545

33. Baar K, Wende AR, Jones TE, et al. Adaptations of skeletal muscle to exercise: rapid increase in the transcriptional coactivator PGC-1. FASEB J. 2002;16(14):1879–1886. doi:10.1096/fj.02-0367com

34. Cartoni R, Léger B, Hock MB, et al. Mitofusins 1/2 and ERRα expression are increased in human skeletal muscle after physical exercise. J Physiol. 2005;567(Pt 1):349–358. doi:10.1113/jphysiol.2005.092031

35. Hecksteden A, Hoppstädter J, Bizjak DA, et al. Effects of acute exercise and training status on glucocorticoid-induced leucine zipper (GILZ) expression in human skeletal muscle. Journal of Science and Medicine in Sport. 2023;26(12):707–710. doi:10.1016/j.jsams.2023.10.007

36. Pelliccia A, Sharma S, Gati S, et al. 2020 ESC Guidelines on sports cardiology and exercise in patients with cardiovascular disease. Eur Heart J. 2021;42(1):17–96. doi:10.1093/eurheartj/ehaa605

37. MacNeil LG, Melov S, Hubbard AE, Baker SK, Tarnopolsky MA. Eccentric exercise activates novel transcriptional regulation of hypertrophic signaling pathways not affected by hormone changes. PLoS One. 2010;5(5):e10695. doi:10.1371/journal.pone.0010695

38. Hyldahl RD, Xin L, Hubal MJ, Moeckel-Cole S, Chipkin S, Clarkson PM. Activation of nuclear factor-κPB following muscle eccentric contractions in humans is localized primarily to skeletal muscle-residing pericytes. FASEB j. 2011;25(9):2956–2966. doi:10.1096/fj.10-177105

39. Zambon AC, McDearmon EL, Salomonis N, et al. Time- and exercise-dependent gene regulation in human skeletal muscle. Genome Biology. 2003;4(10):R61. doi:10.1186/gb-2003-4-10-r61

40. Popov DV, Makhnovskii PA, Shagimardanova EI, et al. Contractile activity-specific transcriptome response to acute endurance exercise and training in human skeletal muscle. American Journal of Physiology-Endocrinology and Metabolism. 2019;316(4):E605–E614. doi:10.1152/ajpendo.00449.2018

41. Page MJ, McKenzie JE, Bossuyt PM, et al. The PRISMA 2020 statement: an updated guideline for reporting systematic reviews. BMJ. 2021;372:n71. doi:10.1136/bmj.n71

42. Edman S, Jones RG, Jannig PR, et al. The 24-Hour Time Course of Integrated Molecular Responses to Resistance Exercise in Human Skeletal Muscle Implicates MYC as a Hypertrophic Regulator That is Sufficient for Growth. Published online March 27, 2024:2024.03.26.586857. doi:10.1101/2024.03.26.586857

43. Louis E, Raue U, Yang Y, Jemiolo B, Trappe S. Time course of proteolytic, cytokine, and myostatin gene expression after acute exercise in human skeletal muscle. Journal of Applied Physiology. 2007;103(5):1744–1751. doi:10.1152/japplphysiol.00679.2007

44. Kostek MC, Chen YW, Cuthbertson DJ, et al. Gene expression responses over 24 h to lengthening and shortening contractions in human muscle: major changes in CSRP3, MUSTN1, SIX1, and FBXO32. Physiological Genomics. 2007;31(1):42–52. doi:10.1152/physiolgenomics.00151.2006

45. Raue U, Trappe TA, Estrem ST, et al. Transcriptome signature of resistance exercise adaptations: mixed muscle and fiber type specific profiles in young and old adults. Journal of Applied Physiology. 2012;112(10):1625–1636. doi:10.1152/japplphysiol.00435.2011

46. McIntosh MC, Sexton CL, Godwin JS, et al. Different Resistance Exercise Loading Paradigms Similarly Affect Skeletal Muscle Gene Expression Patterns of Myostatin-Related Targets and mTORC1 Signaling Markers. Cells. 2023;12(6):898. doi:10.3390/cells12060898

47. R Core Team. R: A Language and Environment for Statistical Computing. Published online 2023.

48. RStudio Team. RStudio: integrated develeopment environment for R. Published online 2023. Accessed April 15, 2024. http://www.rstudio.com/

49. de Torrenté L, Zimmerman S, Suzuki M, Christopeit M, Greally JM, Mar JC. The shape of gene expression distributions matter: how incorporating distribution shape improves the interpretation of cancer transcriptomic data. BMC Bioinformatics. 2020;21(21):562. doi:10.1186/s12859-020-03892-w

50. Terry EE, Zhang X, Hoffmann C, et al. Transcriptional profiling reveals extraordinary diversity among skeletal muscle tissues. Elife. 2018;7:e34613. doi:10.7554/eLife.34613

51. Cowley MJ, Cotsapas CJ, Williams RBH, et al. Intra- and inter-individual genetic differences in gene expression. Mamm Genome. 2009;20(5):281–295. doi:10.1007/s00335-009-9181-x

52. Liu G, Fekete G, Yang H, et al. Comparative 3-dimensional kinematic analysis of snatch technique between top-elite and sub-elite male weightlifters in 69-kg category. Heliyon. 2018;4(7):e00658. doi:10.1016/j.heliyon.2018.e00658

53. Vissing K, Schjerling P. Simplified data access on human skeletal muscle transcriptome responses to differentiated exercise. Sci Data. 2014;1(1):140041. doi:10.1038/sdata.2014.41

54. Deschenes MR, Kraemer WJ. Performance and Physiologic Adaptations to Resistance Training. American Journal of Physical Medicine & Rehabilitation. 2002;81(11):S3.

55. Coffey VG, Shield A, Canny BJ, Carey KA, Cameron-Smith D, Hawley JA. Interaction of contractile activity and training history on mRNA abundance in skeletal muscle from trained athletes. Am J Physiol Endocrinol Metab. 2006;290(5):E849–855. doi:10.1152/ajpendo.00299.2005

56. Schwalm C, Jamart C, Benoit N, et al. Activation of autophagy in human skeletal muscle is dependent on exercise intensity and AMPK activation. FASEB J. 2015;29(8):3515–3526. doi:10.1096/fj.14-267187

57. Yang Y, Jemiolo B, Trappe S. Proteolytic mRNA expression in response to acute resistance exercise in human single skeletal muscle fibers. J Appl Physiol (1985). 2006;101(5):1442–1450. doi:10.1152/japplphysiol.00438.2006

58. Nedergaard A, Vissing K, Overgaard K, Kjaer M, Schjerling P. Expression patterns of atrogenic and ubiquitin proteasome component genes with exercise: effect of different loading patterns and repeated exercise bouts. Journal of Applied Physiology. 2007;103(5):1513–1522. doi:10.1152/japplphysiol.01445.2006

59. Stefanetti RJ, Lamon S, Wallace M, Vendelbo MH, Russell AP, Vissing K. Regulation of ubiquitin proteasome pathway molecular markers in response to endurance and resistance exercise and training. Pflugers Arch - Eur J Physiol. 2015;467(7):1523–1537. doi:10.1007/s00424-014-1587-y

60. Arany Z. PGC-1 coactivators and skeletal muscle adaptations in health and disease. Curr Opin Genet Dev. 2008;18(5):426–434. doi:10.1016/j.gde.2008.07.018

61. Norrbom J, Sundberg CJ, Ameln H, Kraus WE, Jansson E, Gustafsson T. PGC-1α mRNA expression is influenced by metabolic perturbation in exercising human skeletal muscle. Journal of Applied Physiology. 2004;96(1):189–194. doi:10.1152/japplphysiol.00765.2003

62. Takahashi A, Honda Y, Tanaka N, et al. Skeletal Muscle Electrical Stimulation Prevents Progression of Disuse Muscle Atrophy via Forkhead Box O Dynamics Mediated by Phosphorylated Protein Kinase B and Peroxisome Proliferator-Activated Receptor gamma Coactivator-1alpha. Physiol Res. 2024;73(1):105–115. doi:10.33549/physiolres.935157

63. Koh JH, Pataky MW, Dasari S, et al. Enhancement of anaerobic glycolysis – a role of PGC-1α4 in resistance exercise. Nat Commun. 2022;13(1):2324. doi:10.1038/s41467-022-30056-6

64. Ruas JL, White JP, Rao RR, et al. A PGC-1α isoform induced by resistance training regulates skeletal muscle hypertrophy. Cell. 2012;151(6):1319–1331. doi:10.1016/j.cell.2012.10.050

65. Centner C, Jerger S, Mallard A, et al. Supplementation of Specific Collagen Peptides Following High-Load Resistance Exercise Upregulates Gene Expression in Pathways Involved in Skeletal Muscle Signal Transduction. Front Physiol. 2022;13:838004. doi:10.3389/fphys.2022.838004

66. Dickinson JM, D’Lugos AC, Naymik MA, et al. Transcriptome response of human skeletal muscle to divergent exercise stimuli. Journal of Applied Physiology. 2018;124(6):1529–1540. doi:10.1152/japplphysiol.00014.2018

67. Makhnovskii PA, Gusev OA, Bokov RO, et al. Alternative transcription start sites contribute to acute-stress-induced transcriptome response in human skeletal muscle. Hum Genomics. 2022;16(1):24. doi:10.1186/s40246-022-00399-8

68. Popov DV, Makhnovskii PA, Kurochkina NS, Lysenko EA, Vepkhvadze TF, Vinogradova OL. Intensity-dependent gene expression after aerobic exercise in endurance-trained skeletal muscle. Biol Sport. 2018;35(3):277–289. doi:10.5114/biolsport.2018.77828

69. Rowlands DS, Thomson JS, Timmons BW, et al. Transcriptome and translational signaling following endurance exercise in trained skeletal muscle: impact of dietary protein. Physiological Genomics. 2011;43(17):1004–1020. doi:10.1152/physiolgenomics.00073.2011

70. Neubauer O, Sabapathy S, Lazarus R, et al. Transcriptome analysis of neutrophils after endurance exercise reveals novel signaling mechanisms in the immune response to physiological stress. Journal of Applied Physiology. 2013;114(12):1677–1688. doi:10.1152/japplphysiol.00143.2013

71. Rubenstein AB, Hinkley JM, Nair VD, et al. Skeletal muscle transcriptome response to a bout of endurance exercise in physically active and sedentary older adults. Am J Physiol Endocrinol Metab. 2022;322(3):E260–E277. doi:10.1152/ajpendo.00378.2021

