## Supplemental Data Acquisition for "Resistance but not endurance training suppresses glucocorticoid-induced leucine zipper (GILZ) expression in human skeletal muscle"

**Supplementary materials**

*2 METHODS*

*2.1 Data acquisition*

*2.1.1 Query strings*

The Gene Expression Omnibus (GEO) Repository was searched exclusively. The query for data has been subdivided based on the implemented exercise regime. Following query strings were used:

- Query string for endurance exercise: *human[organism] AND (endurance training OR aerobic training OR "endurance exercise" OR "endurance training") AND ("expression profiling by array"[DataSet Type] OR “Expression profiling by high throughput sequencing”[DataSet Type])*
- Query string for resistance exercise (traditional and eccentric): *human[organism] AND (resistance training OR strength training OR resistance exercise) AND ("expression profiling by array"[DataSet Type] OR “Expression profiling by high throughput sequencing”[DataSet Type])*

The search ended on February 29, 2024.

*2.2.2 Identification of transcript/gene probes on micro array/high-throughput sequencing platforms*

Table 1 lists the IDs that were used to identify the probes, corresponding to the gene of interest, on the respective microarray or high-throughput sequencing platform. Generally, if the search for the respective RefSeq ID did not yield any results, the gene name, the Ensembl ID, and the alternative search term were used, in this order. It is noteworthy that in one case the atrogenes MuRF1 and MAFbx were not found on platform GPL8300, used by Zambon and coworkers[1].The same is true for PGC1$\alpha$, which was also not found on platform GPL8300.

*Table 1: Search terms used to identify corresponding microarray/high-throughput sequencing probe.*

| Gene of interest | Gene name | RefSeq ID | Ensembl ID | Alternative |
|---|---|---|---|---|
| GILZ | TSC22D3 | NM_004089 | ENST00000372397 | / |
| MAFbx | FBXO32 | NM_058229 | ENST00000517956 | Atrogin1 |
| MuRF1 | TRIM63 | NM_032588 | ENST00000374272 | / |
| Cathepsin-L | CTSL | NM_001912 | ENST00000343150 | / |
| PGC1$\alpha$ | PPARGC1A | NM_013261 | ENST00000264867 | / |

REFERENCE

1. Zambon AC, McDearmon EL, Salomonis N, et al. Time- and exercise-dependent gene regulation in human skeletal muscle. *Genome Biology*. 2003;4(10):R61. doi:10.1186/gb-2003-4-10-r61
